# Multiplex Fragment Analysis Identifies SARS-CoV-2 Variants

**DOI:** 10.1101/2021.04.15.21253747

**Authors:** A.E. Clark, Z. Wang, B. Cantarel, M. Kanchwala, C. Xing, L. Chen, P. Irwin, Y. Xu, D. Oliver, F. Lee, J.R. Gagan, L. Filkins, A. Muthukumar, J.Y. Park, R. Sarode, J.A. SoRelle

## Abstract

The rapid spread of SARS-CoV-2 Variants of Concern (VOC) necessitates systematic efforts for epidemiological surveillance. The current method for identifying variants is viral whole genome sequencing (WGS). Broad clinical adoption of sequencing is limited by costly equipment, bioinformatics support, technical expertise, and time for implementation. Here we describe a scalable, multiplex, non-sequencing-based capillary electrophoresis assay to affordably screen for SARS-CoV-2 VOC.

## Main

Novel strains of SARS-CoV-2 have emerged with clinically significant genetic mutations. These “variants of concern” VOCs impact transmission kinetics,^1^ vaccine responses,^2,3^ mortality,^4,5^ and drug resistance (monoclonal antibodies).^6–9^ Therefore, methods for real-time identification of SARS-CoV-2 VOCs are of exigent need in both clinical and public health arenas. Whole-genome sequencing (WGS) is the current gold standard for SARS-CoV-2 variant identification; however, broad WGS adoption is hampered by cost and requirements for specialized equipment and bioinformatics expertise.^10^ While these limitations may be mitigated through centralized high-volume testing, the consequent increased turnaround time for batching, data deconvolution, and sequence analysis exceeds what is clinically actionable for most patient assessments. Therefore, simple, on-site, positive specimen reflex testing for VOCs represents a practical solution.

Here, we describe a rapid, cost-effective, and scalable method to detect known VOCs utilizing fragment analysis by capillary electrophoresis in a routine clinical setting. In this procedure, extracted RNA is amplified to create fluorescently-labeled amplicons separated by capillary electrophoresis on a Sanger sequencer. VOCs are then identified by their abnormal amplicon sizes compared to a wild-type reference. By measuring the resultant changes in fragment length, this assay permits a broader range of variant detection compared to primer-probe based RT-qPCR. The reagent and instrumentation requirements for this assay are modest: fluorescently labeled primers, a thermocycler, and Sanger sequencer. Moreover, the time requirement (4 hours, Fig. 1B), simplicity, and cost-effective use of 96-well format support the assay’s scalability. Amplicons can be designed up to 500bp in length with up to four fluorescent labels, making the assay adaptable to detect additional SARS-CoV-2 variants as they are identified (Fig. 1C, potential to multiplex to 20-40 targets). This assay therefore allows routine clinical laboratories to rapidly screen SARS-CoV-2 positive specimens for VOCs by a mechanism that can be easily and broadly implemented with limited financial burden or bioinformatic expertise. Real-time monitoring of SARS-CoV-2 VOCs could impact epidemiological tracking and guide clinical management prior to administering monoclonal antibody therapy.

**Figure 1.**
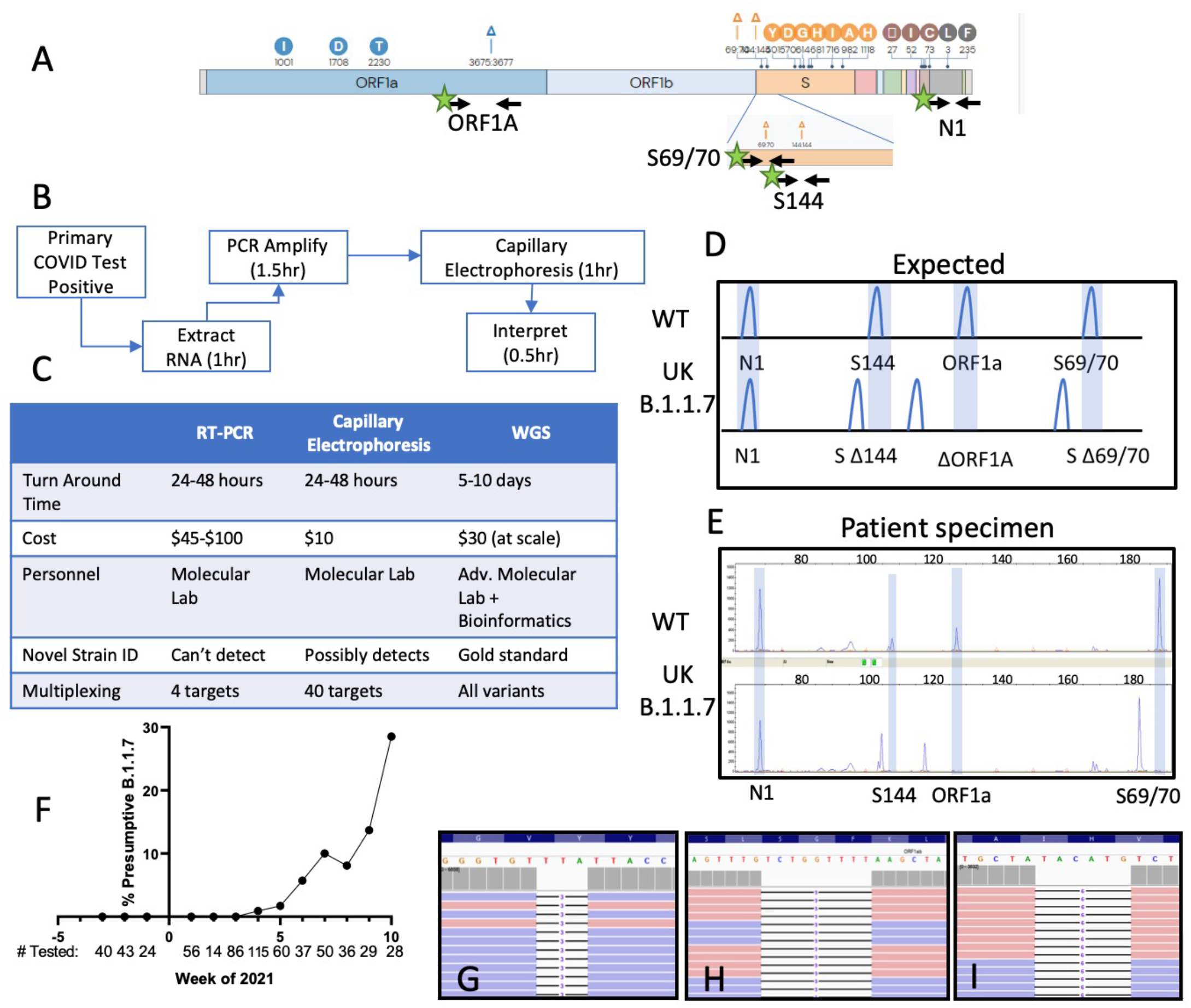
Fragment analysis identifies the SARS-CoV-2 B.1.1.7 Variant of Concern. Genetic variations of the SARS-CoV-2 B.1.1.7 (UK) VOC. Black arrows indicate primer binding sites. Green stars indicating fluorescent dye (FAM) primer labels. (B) Routine clinical workflow and timetable of the SARS-CoV-2 fragment analysis assay. (C) Comparison of analytical and operational characteristics of real time PCR (RT-qPCR), fragment analysis by capillary electrophoresis and WGS (whole genome sequencing). (D) Expected deletions present in the SARS-CoV-2 B.1.1.7 strain compared to wild type as visualized by fragment analysis. (E) Electrophoretogram representative of two patients with WT and B.1.1.7 SARS-CoV-2 infections. (F) Estimated prevalence of presumptive B.1.1.7 per week with number of SARS-CoV-2 positive specimens tested below. (G-I) Integrated genome viewer visualization of the whole genome sequencing reads confirming the presence of the ORF1A:3675_3677del (G), S:69_70del (H), and S:144del (I) initially identified by capillary electrophoresis.

For proof of concept, we targeted three deletion mutations initially described in the UK B.1.1.7 VOC. The B.1.1.7 strain was originally discovered due to S-gene target failure (SGTF) secondary to the Spike gene HV69_70del variant in a SARS-CoV-2 commercial assay.^1,5,11^ While present in B.1.1.7, the HV69_70del is not specific to this strain (55% of northeast U.S. HV69_70del mutants are not B.1.1.7);^12^ therefore, we targeted the S-gene F144 deletion mutation, to improve specificity for B.1.1.7 to >99%.^13^ Frequent in-frame deletions near the 69/70 and 144 amino acid positions suggest this is a mutational hotspot, which could arise in other emerging strains.^14^ An ORF1A deletion (SDF3675_3677del) is present in B.1.1.7 (UK variant),^4^ B.1.351 (South African, originally N150Y.2),^15^ P.1 (Brazil),^16^ and B.1.526 (New York) variants. Therefore, detecting an ORF1A deletion without S gene deletions is suggestive of the B.1.351, P.1, or B.1.526 VOCs.^17^ (Fig. 1A, 2G). Lastly, the CDC N1 primer pair was added as an internal positive control.

We validated this method focusing on the principle SARS-CoV-2 VOC impacting European countries and the United States: B.1.1.7.^11^ An initial screen of 182 retrospective (12/1/20-1/20/21) and 466 prospective SARS-CoV-2 positive nasopharyngeal specimens (1/25/21-3/11/21) was performed. No VOCs were identified among retrospective specimens (0/182). On the third week in January 2021, the first VOC was identified with characteristic shortened amplicons reflecting B.1.1.7 deletions (Fig 1D, E, Interpretive criteria in Supplemental Methods). This deletion pattern was confirmed in control RNA containing the B.1.1.7 sequence. For verification, the suspected B.1.1.7 isolate and positive control sample were sequenced by WGS (Supplemental methods), and classified by PANGOLIN as belonging to the B.1.1.7 lineage (Fig. 1G-I).^18^ 

Overall, we detected 32 specimens containing SARS-CoV-2 consistent with B.1.1.7 (n=23 patient specimens, n=9 control RNA, 31 WGS verified, 1 apparent false positive). All specimens had low SARS-CoV-2 RT-PCR CT values (CT <30), and each displayed an identical deletion pattern by fragment analysis. Strikingly, B.1.1.7 was absent in our patient population before January 2021, but prevalence has steadily increased each week. Using this method, we estimate the weekly prevalence rate to be 28.5% (8/28) in the last week of the study (3/8/21-3/12/21, Fig. 1F) despite declining rates of COVID-19 cases in north Texas. This is consistent with the trend currently observed in the United Kingdom.^19^ 

During validation studies, we realized the reverse primer for S144 overlapped with the W152C mutation present in the emerging B.1.429/ B.1.427 variants (California, CAL.20C).^20^ The primer was thus redesigned to bind the wild type position preferentially and fail to amplify when the S:W152C (22018G>T) variant occurs. To further destabilize S144R binding to the 22018G>T variant, nucleotides at the 2^nd^and 3^rd^positions upstream from 3’ terminus were mutated. (Fig. 2C, D). Primer S144_R2’T>G was tested against 4 WGS confirmed B.1.429 variants with W152C mutations (Fig. 2H). However, S144 dropout was not detected for any primer pair. Instead, we detected a single base pair deletion (Fig. 2A, B). This was clearest in the primer with the 2^nd^upstream nucleotide mutated from T>G (Supplemental Table 1, S144_R2’T>G). An alternative primer binding configuration skipping the 22018G>T variant could explain the 1 base pair deletion. This aberration provides specificity without loss in sensitivity for California variants, thus demonstrating the assay’s flexibility to adapt to emerging strains.

**Figure 2.**
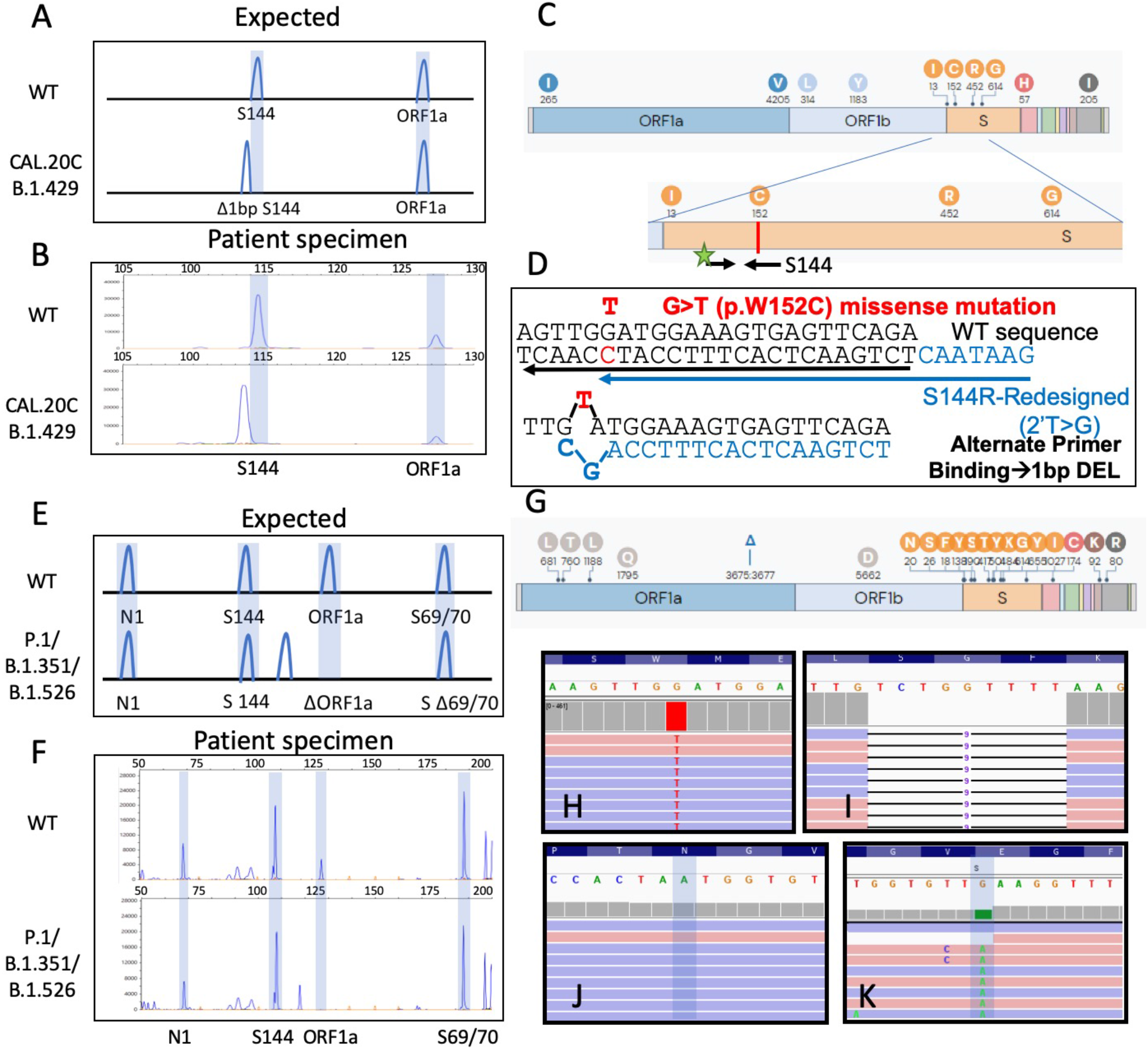
Detection of other SARS-CoV-2 variants by fragment analysis. (A) Expected deletions present in B.1.429 (California) strain compared to wild type SARS-CoV-2. (B) Electrophoretogram representative of two patients with wild-type and B.1.429 SARS-CoV-2 infections. (C) Genetic variation of the SARS-CoV-2 B.1.429 strain. Black arrows indicate the primer binding sites for S144 target primers. The red line indicates the location of the W152C genetic variant overlapping with the original S114 reverse primer. (D) Detailed view of the variant causing W152C and proposed alternative primer binding site of S114 redesigned primer (blue arrow and blue text), which accounts for the single nucleotide deletion observed. (E) Expected deletions present in P.1, B.1.351, and B.1.526 strains compared to WT (wild type) SARS-CoV-2. (F) Electrophoretograms representative of two patients: one with wild type SARS-CoV-2 infection, the second with a virus that exhibits a nine base pair deletion in ORF1A. (G) Genetic variation of the SARS-CoV-2 P.1 strain. (H) Integrated genome viewer visualization of the whole genome sequencing reads confirming the presence of the SW152C variant characteristic of the B.1.429 strain (H) and ORF1A:3675_3677del (I), which is found in each Variant of Concern. Lack of the N501Y (J) and E484K (K) mutations precludes classification of this SARS-CoV-2 strain as P.1/ B.1.351.

When the assay was designed, an isolated ORF1A:del3675_3677 (without S gene deletions) was thought to differentiate P.1 and B.1.351 variants from B.1.1.7 (Fig. 2E, G). Therefore, when we detected isolated 9 nucleotide deletions in the ORF1A target, these VOCs were suspected (n=5). Interestingly, although WGS confirmed the ORF1A:3675_3677 deletion (Fig. 2I), neither P.1 nor B.1.351 were identified. Rather, three strains were classified as B.1.526 (N501Y absent and E484K present, Fig.2 J,K) and the other two cases as B.1.241 (N501Y and E484K absent). This recurrence of ORF1A:del3675_3677 in multiple VOC indicates ORF1A:DEL3675_3677 may represent convergent evolution arising in geographically disparate locations. This assay can therefore detect mutations consistent with clinically important SARS-CoV-2 strains, as well as monitor for emerging strains with related genetic alterations.

The limit of detection varies by target but ranges from 2,000 copies/ reaction for ORF1A to 62.5 copies/ reaction for all additional targets (Supplemental Table 5). Furthermore, as contamination is a concern for PCR reactions with high levels of amplification, this assay can detect the presence of a mixture of WT and mutant DNA by identifying the existence of multiple peaks.

Epidemiologic data indicates co-occurrence of the three deletions targeted are present in 95% of B.1.1.7 strains. Thus, fragment analysis of these three loci may be sufficient for B.1.1.7 classification. A similar principle may be true for other strains with unique genetic mutations. Additionally, the high rate of mutational change within the SARS-CoV-2 spike protein may lead to additional serendipitous findings, similar to the original SGTF.

In summary, we have applied fragment analysis to SARS-CoV-2 VOC detection and epidemiology for the first time. This assay can detect characteristic viral genomic changes found in B.1.1.7 (UK), P.1 (Brazil), B.1.351 (South African), B.1.529 (New York), and B.1.427/ B.1.429 (California) lineages. This approach is cost-effective, multiplexable (up to 20-40 primer pairs can be added), scalable (up to 96 specimens), and easily adaptable to routine clinical workflow in laboratories with molecular biology expertise and instrumentation. Our future directions include adding additional characteristic markers of P.1/ B.1.351/ B.1.526/ B.1.429 (i.e. S:N501Y, S:E484K, S:242_244del, S:L452R, and Ins28269_28273) to distinguish strains with 95% accuracy.^21^

## Data Availability

Data are available upon request provided it does not interfere with publication in a peer-review journal.

## Acknowledgements

The authors gratefully acknowledge the technical assistance of Hui Zheng, Huiyu Yao, Emily Ostman, Vanessa Torres and Ginger Koetting. Additionally, we thank Prithvi Raj and Vanessa Schmid for sequencing assistance, and all members of the UTSW COVID diagnostic laboratory for archiving and retrieving specimens.

