## Supplemental Material and Methods for "Multiplex Fragment Analysis Identifies SARS-CoV-2 Variants"

**SARS-CoV-2 variant control material.** Positive control material for all B.1.1.7 variants consisted of commercially ordered synthetic RNA (Synthetic SARS-CoV-2 RNA Control 14, GISAID name: England/205041766/2020) produced as six, 5kb fragments (Twist Bioscience, San Francisco, CA).

**Clinical specimens.** Clinical specimens from the pathology service line at the University of Texas Southwestern Medical Center were utilized for assay verification. 182 retrospective (12/1/20-1/20/21) and 466 prospective SARS-CoV-2 specimens (1/25/21-3/7/21) were examined. The testing indication ranged from asymptomatic pre-procedure screening to symptomatic and previously confirmed COVID-19 cases. Samples consisted of nasopharyngeal swabs in viral transport media (Remel, ThermoScientific, Lenexa, KS) which were previously determined to contain SARS-CoV-2 in the routine setting by either RT-PCR (RealTi*m*e SARS-CoV-2/*m*2000, Alinity *m* SARS-CoV-2, Abbott Molecular, Des Plaines, IL) or isothermal amplification (IDNow, Abbot Diagnostics, Scarbourough, ME). Included specimens had qRT-PCR CN values of <30 (Alinity m) or <20 (m2000 platform) to assure enough material for analysis. Positive specimens not meeting these criteria were excluded.

**Oligonucleotides.** Oligonucleotide primer sequences were based on previously published designs (N1, S144, ORF1A, and S69/70) and can be found in Table 1.[1–3] All primers were ordered as desalted and dried oligonucleotides from either Integrated DNA Technologies (IDT, Coralville, IA) or Eurofins (Louisville, KY). All primers were resuspended in IDTE buffer or nuclease free water (ThermoFisher Scientific, Waltham MA) to a final concentration of 100µM prior to use. Primers were designed and analyzed using the Multiple Primer Analyzer webtool by ThermoFisher Scientific (https://www.thermofisher.com/us/en/home/brands/thermo-scientific/molecular-biology/molecular-biology-learning-center/molecular-biology-resource-library/thermo-scientific-web-tools/multiple-primer-analyzer.html). A FAM label was added to the 5' end of all forward primers.

Table. 1

| Name | Sequence | Tm°C | CG% | nt | Position | Amino Acids |
|---|---|---|---|---|---|---|
| S69/70_F: | 5'-FAM-cgtggtgtttattaccctgacaaag-3' | 66.3 | 44 | 25 | c.21611-21792 | 34-94 |
| S69/70_R: | 5'-tcagtggaagcaaaataaacaccat-3' | 66.5 | 36 | 25 | | |

| | | | | | | |
|---|---|---|---|---|---|---|
| S144_F: | 5’-FAM-acgctactaatgttgttattaaagtct-3’ | 59 | 34.5 | 27 | c.21912-22018 | 120-155 |
| S144_R: | 5’-tctgaactcactttccatccaact-3’ | 65.2 | 41.7 | 24 | | |
| S144_R2’T>G: | 5’- gaataactctgaactcactttccagc-3’ | 65.2 | 41.7 | 24 | | |
| ORF1A_F: | 5’-FAM-tgcctgctagttgggtgatg-3’ | 66.5 | 55 | 20 | c.11229-11347 | N/A |
| ORF1A_R: | 5’-tgctgtcataaggattagtaacact-3’ | 60.1 | 36 | 25 | | |
| N1_F: | 5’-FAM-gaccccaaaatcagcgaaat-3’ | 65.2 | 45 | 20 | c.28268-28339 | 5-29 |
| N1_R: | 5’-tctggttactgccagttgaatctg-3’ | 66.5 | 45.8 | 24 | | |

**RNA Extraction.**

RNA extraction was performed using chemagic™ Viral DNA/RNA 300 Kit H96 executed on chemagic™ 360 instrument (PerkinElmer, Hopkinton, MA) according to the manufacturer’s protocol. A sample plate, an elution plate and a magnetic bead plate were prepared using an automated liquid handling instrument (Janus G3 workstation, PerkinElmer Inc). In brief, an aliquot of 300µl from each sample, 4µL Poly(A) RNA, 10µL proteinase K and 300µL lysis buffer 1 were added to respective wells of a 96 well sample plate. The sample plate, elution plate (60µL elution buffer per well) and magnetic beads plate (150µL beads per well) were then placed on the chemagic™ 360 instrument and RNA was extracted automatically with an elution volume of 60µL from a sample volume of 300 µL.

**RT-PCR Amplification.** Prior to RT-PCR, all reagents were thawed on ice, gently vortexed and centrifuged to assure homogeny. A 10X RT-qPCR primer master mix was created using 5µL of the 100µM stock of each primer from each of the 8 unique primers described in Table 1 added into 455µL of nuclease-free water to total 500µL. RT-PCR was performed using the Thermo TaqPath 1-Step RT-qPCR kit (ThermoFisher Scientific, Waltham MA). Briefly, 2.5µL 4X TaqPath master mix (including M-MLV reverse transcriptase and DNA polymerase), 1.5µL nuclease-free water, and 5µL extracted RNA were added per reaction. 1µL of the 10X primer master mix was added per reaction, resulting in a total volume of 10µL per reaction. This mixture was scaled to either single, 50 or 96 reactions depending on the application (see Table 2). Single reactions were performed in standard PCR tubes, while group of 96 reactions were performed utilizing a 96-well plate. RT-PCR amplification was performed using a modification of the thermocycler settings for the CDC SARS-CoV-2 assay[4] to include a 30-

second extension step at 72°C to account for larger amplification fragments generated in this application compared to traditional RT-qPCR.

The RT-qPCR cycling program used for the assay is as follows: 1. *Initial denaturation:* 2 minutes at 25°C, 15 minutes at 50°C, 2 minutes at 95°C, 2. *Amplification:* 40 cycles of: 95°C for 10 seconds, 60°C for 30 seconds, 72°C for 30 seconds, 3. *Final extension:* 72°C for 5 minutes, terminal hold at 4°C. Following amplification, RT-PCR products were either immediately analyzed by capillary electrophoresis, or alternatively were stored at -20°C protected from light until analysis could be undertaken.

**Capillary Electrophoresis.** Following RT-PCR, 8.5µL Hi-Di Formamide (ThermoFisher) and 0.5µL of GeneScan™ 500 LIZ™ standard (ThermoFisher) was added to each well of a new 96-well plate, or a fresh tube if fewer specimens were analyzed. 1µL of RT-PCR product was added to each well, and 96-well plates were sealed with adhesive film. Tubes or plates were briefly vortexed and centrifuged gently to assure a homogenous mixture, and then amplification fragments were heat denatured at 95°C for 3 minutes. Samples were then returned to ice and protected from light. The Sanger sequencer (Applied Biosystems 3730xl, 50cm capillaries, Polymer POP-7r) was calibrated for LIS and FAM dyes prior to running samples, and adhesive film was removed from the 96-well plate containing the denatured RT-PCR products.

**Interpretive criteria.** The expected wild-type and SARS-CoV-2 variant fragment lengths were confirmed (Table 2). The N1 target served as a positive control for the presence of SARS-CoV-2. Samples were considered positive if capillary electrophoresis peaks were: 1) of the expected size (x-axis, Table 2) and 2) >100 fluorescence units. The cut-off of 100 fluorescence units was determined as being 3x higher than baseline fluorescence for the respective fluorescence channel. A small number of recurrent artifacts were observed, and are described in Table 3. Considering all targets together can lead to presumptive classification of variants as either wild type, B.1.1.7, B.1.351/P.1, B.1.429 (Table 4).

Table 2. Expected WT and mutant amplicon size

| Target | Mutation | Base pairs deleted | WT amplicon size | Mutant amplicon size |
|---|---|---|---|---|
| **S69_70del** | 21765_21770del | 6 bp | 188 bp | 182 bp |
| **S144del** | 21991_21993del | 3 bp | 107 bp | 104 bp |
| **ORFA1Adel** | 11288_11296del | 9 bp | 127 bp | 118 bp |
| **CDC_N1** | N/A | N/A | 68 bp | N/A |

Table 3. Recurrent Artifacts

| Location | Comments | Location | Comments |
|---|---|---|---|
| **<50** | Primer dimers | 96.8 | Broad |
| **87.8** | Broad | 162bp | Broad |
| **91.2** | Broad | | |

Table 4. SARS-CoV-2 Variant Interpretation

| Targets | Wild Type | Presumptive B.1.1.7 | Potential B.1.351 or P.1 | Presumptive B.1.429 |
|---|---|---|---|---|
| **S69_70del** | WT | 6bp deletion | WT | WT |
| **S144del** | WT | 3bp deletion | WT | 3bp insertion |
| **ORF1Adel** | WT | 9bp deletion | 9 bp deletion | WT |
| **N1** | WT | WT | WT | WT |

**Limit of Detection**

The limit of detection was estimated using spiked-in B.1.1.7 in vitro transcribed RNA in triplicate. The number of copies per reaction ranged from 20,000 to 62.5 copies, and positivity per target was displayed below.

Table 5. Limit of detection study with B.1.1.7 RNA.

| Targets | N1 | S144 | ORF1a | S69/70 |
|---|---|---|---|---|
| **20,000** | 3/3 | 3/3 | 3/3 | 3/3 |
| **4,000** | 3/3 | 3/3 | 3/3 | 3/3 |
| **2,000** | 3/3 | 3/3 | 3/3 | 3/3 |
| **1,000** | 3/3 | 3/3 | 0/3 | 3/3 |
| **500** | 3/3 | 3/3 | 1/3 | 3/3 |
| **250** | 3/3 | 3/3 | 0/3 | 3/3 |
| **125** | 3/3 | 3/3 | 0/3 | 3/3 |
| **62.5** | 3/3 | 3/3 | 0/3 | 3/3 |

**Next generation sequencing.**

First strand cDNA was prepared using 10uL of RNA according to the manufacturer's instructions (Prime Script 1st strand cDNA synthesis, Takara Bio, Japan). Five microliters of cDNA was converted into an NGS library with the Swift SNAP SARS-CoV-2 Kit (https://swiftbiosci.com/wp-content/uploads/2021/01/PRT-028-Swift-Normalase-Amplicon-Panels-SNAP-SARS-CoV-2-Panels-Rev-6.pdf) using tiled primer pairs in a single tube to target the 29.9 kb viral genome. Samples were sequenced on the Illumina® MiSeq® System at 2 x 250 bp. Sequencing primers were designed against the NCBI Reference Sequence NC_045512.2 (Severe acute respiratory syndrome coronavirus 2 isolate Wuhan-Hu-1, complete genome).

**Bioinformatic Analysis.**

Raw sequences were trimmed for quality and adapter sequence using TrimGalore (https://github.com/FelixKrueger/TrimGalore) with a quality threshold of 25. Trimmed reads were aligned with minimap2[5] to the SARS-COV-2 reference genome (NC_045512). Primer sequences were removed using primerclip (https://github.com/swiftbiosciences/primerclip) Alignments were sorted and indexed using samtools.[6] Duplicate reads were removed using picard MarkDuplicates (https://broadinstitute.github.io/picard/). Variants were detected using BCFtools and VarScan.[7] Consensus sequences were determined using BCFtools. Variants were annotated using snpEFF.[8] Strain lineage was determined using PANGOLIN.[9]
